# Preliminary evaluation of voluntary event cancellation as a countermeasure against the COVID-19 outbreak in Japan as of 11 March, 2020

**DOI:** 10.1101/2020.03.12.20035220

**Authors:** Yoshiyuki Sugishita, Junko Kurita, Tamie Sugawara, Yasushi Ohkusa

**Author notes:** Corresponding author: Yoshiyuki Sugishita.

## Abstract

**Background:** To control COVID-19 outbreak in Japan, sports and entertainment events were canceled in Japan for two weeks from 26 February to 11 March. It has been designated as voluntary event cancellation (VEC).

**Object:** This study predicts the effectiveness of VEC enduring and after its implementation.

**Method:** We applied a simple susceptible–infected–recovery model to data of patients with symptoms in Japan during 14 January to VEC introduction and after VEC introduction to 8 March. We adjusted the reporting delay in the latest few days.

**Results:** Results suggest that the basic reproduction number, R_0,_ before VEC introduced as 2.50 with a 95% confidence interval (CI) was [2.43, 2.55] and the effective reproduction number, R_v,_ after VEC introduced as 1. 88; its 95% CI was [1.68,2.02].

**Discussion and Conclusion:** Results demonstrated that VEC can reduce COVID-19 infectiousness by 35%, but R_0_ remains higher than one.

## Introduction

The initial case of COVID-19 in Japan was that of a patient returning from Wuhan, China who showed symptoms on 3 January, 2020. Subsequently, as of 11 March, 2020, 503 cases were announced, including asymptomatic or those abroad in countries such as China but excluding those infected on a large cruise ship, the Diamond Princess [1].

Sports and entertainment events were canceled in Japan for two weeks from 26 February to 11 March according to a government advisory. At the same time, it was advised that small business and private meetings be cancelled voluntarily. The policy is designated as voluntary event cancellation (VEC). Moreover, since 3 March, almost all schools have been closed to control the spread until early April. Although schoolchildren are not of pestiferous age, the policy effects remain unknown. These policies must be evaluated as soon as possible. If the effective reproduction number, R_v_ under these measures is less than one, the outbreak can be contained. Alternatively, even given a low R_0_ before these measures, if it was greater than one, it might prolong the outbreak. Nevertheless, one could expect to prevent some fatal cases over time by easing burdens on medical resources or developing a vaccine. The entire course of the outbreak might be altered if these measures were able to reduce infectiousness considerably. Therefore, evaluation of these measures must be thorough when these measures are commenced, continued, and ceased. The present study was conducted to evaluate VEC before VEC is cancelled so as to contribute government’s decision making whether VEC will be continued or ceased.

## Method

We applied a simple susceptible–infected–recovery (SIR) model [2] to the data assuming an incubation period following its empirical distribution in the early stage of the outbreak in Japan. Experiences of Japanese people living in Wuhan until the outbreak provide information related to mild cases because complete laboratory surveillance was administered for them. During January 29 – February 17, 2020, 829 Japanese people returned to Japan from Wuhan. All had received a test to detect COVID-19; of them, 14 were found to be positive for COVID-19 [3]. Of those 14, 10 Japanese people had exhibited mild symptoms; the other 4 showed no symptom as of February 25. Moreover, two Japanese residents of Wuhan exhibited severe symptoms: one was confirmed as COVID-19. The other died, although no fatal case was confirmed as COVID-19 by testing. In addition, two Japanese residents of Wuhan with mild symptoms were refused re-entry to Japan even though they had not been confirmed as infected. If one assumes that the Japanese fatal case in Wuhan and that the two rejected re-entrants were infected with COVID-19, then 2 severe cases, 12 mild cases, and 4 asymptomatic cases were found to exist among these Japanese residents of Wuhan. We therefore apply these proportions to the simulation.

Assuming that the power of infectivity among severe patients and mild patients were equal among the asymptomatic cases, half of the symptomatic cases can be assumed. This assumption about relative infectiousness among asymptomatic cases compared with symptomatic cases was also assumed in simulation studies for influenza [4–8]. We sought R_0_ to fit the number of patients during 14 January – 28 February and to minimize the sum of absolute values of discrepancies among the reported numbers and the fitted values. Its 95% confidence interval (CI) was calculated using the 10000 iterations of bootstrapping for empirical distribution.

We used data of the community outbreak of patients with COVID-19 who showed any symptom in Japan for 14 January – 8 March, 2020. During this period, 432 cases with onset dates occurred. We excluded some patients who had returned from China, and who were presumed to be infected persons from the Diamond Princess. They were presumed to be not community-acquired in Japan.

Published information about COVID-19 patients with symptoms from the Ministry of Labour, Health and Welfare (MLHW), Japan was usually adversely affected with some delay caused by uncertainty during onset to visiting a doctor or in the timing of a physician’s suspicion of COVID-19. Therefore, published data of patients must be adjust at least a few days. To adjust it, we applied the following regression. We denote *Xt-k/t* as the number of patients whose onset date was *t-k* published on day *t*. The dependent variables are the degree of reporting delay, *Xt-k-m/t* / *Xt-k-m/t-m*, where *k*>*m* in several *m* and *k*. Here, *m* denotes the difference of the publishing dates between the two published. Date *t* represents the publishing date of the latest publishing. The explanatory variables were 1/*k*, 1/*m*, and 1/*km*. The degree of reporting delay was estimated as [estimated coefficient of constant term] + [estimated coefficient of 1/*k*]/*k*, when *m* was sufficiently large and time had passed. Therefore, this estimated degree of reporting delay multiplied by the latest published data are expected to be predictions of the number of patients whose onset date was *t*-*k*. We used this adjusted number of patients in the latest few days including those after VEC was adopted. We used the published data on 2,5,6, 9,10, 11 and 12 March,2020 by MLHW[1].

First, we estimated R_0_ in Japan to fit the data of community outbreak before VEC was introduced. Then, using the adjusted data of patients, we estimated R_v_ after VEC was adopted.

## Results

During 14 January – 8 March in Japan, 412 community-acquired cases were identified for whom the onset date was published. Figure 1 showed the empirical distribution of incubation period among 59 cases whose exposed date and onset date were published by MHLW. Its mode was six days and average was 6.6 days.

**Figure 1:**
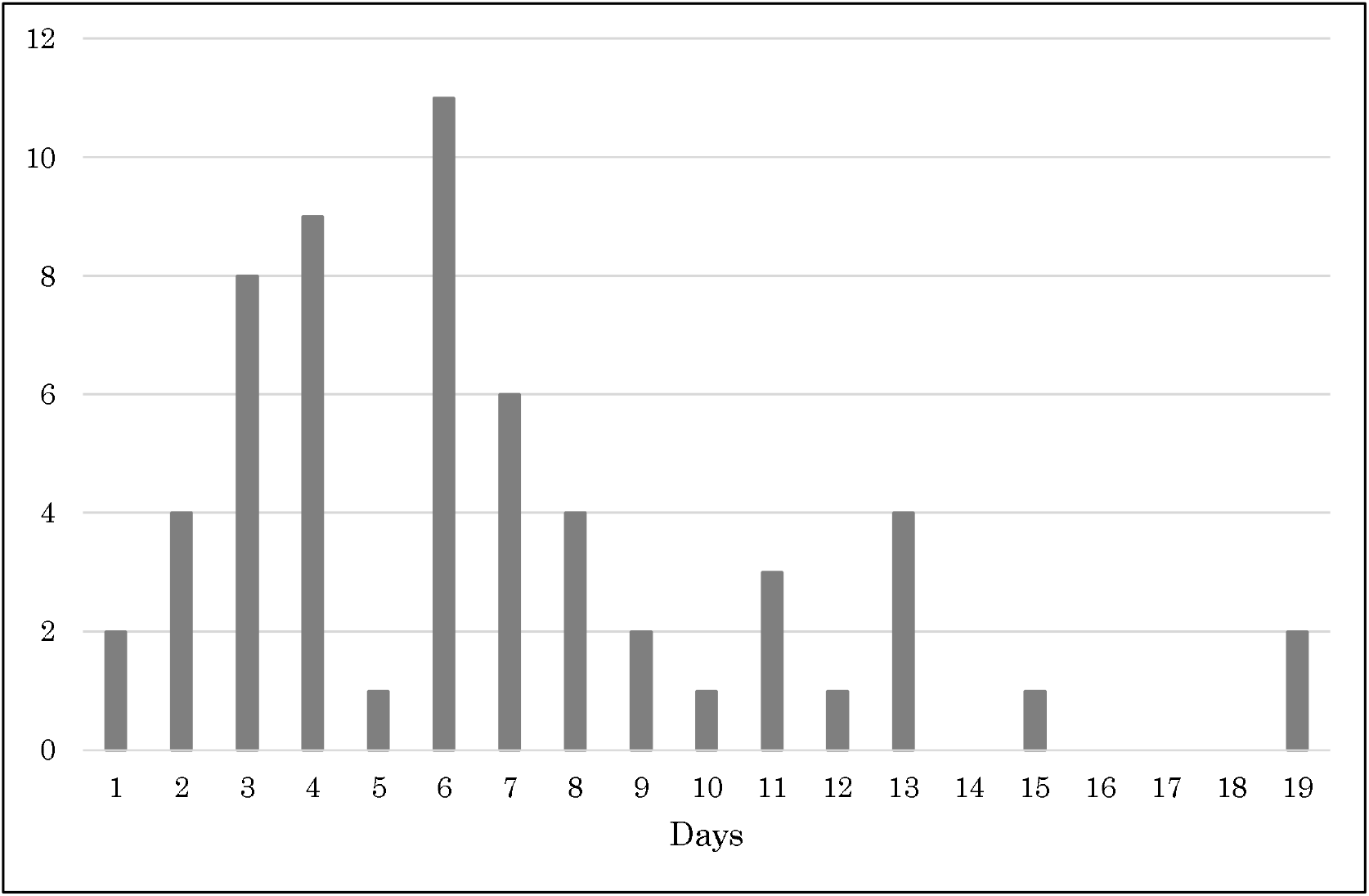
Epirical distributionof incubation period published by Ministry of Labour, Health and Welfare, Japan. (number of patients) Notes: Bars indicates the number of patients by incubation period among 59 cases whose exposed date and onset date were published by Ministry of Labour, Health and Welfare, Japan.

Figure 2 depicts the epidemic curves published at 3, 5, 6, 9 and 10 March. From this information, we estimated the degree of reporting delay. Those results are presented in Table 1. The table shows that 1*/k*, 1/*m* and 1/*km* are all significant. When *m* is sufficiently large, the effects of 1/*m* and 1/*km* converge to zero. Therefore, the estimated degree of reporting delay consists of the term of 1/*k* and a constant term. Based on these results, we predict the degrees of reporting delay as 17.3 for *k*=1, 8.72 for *k*=2, 5.84 for *k*=3, and so on.

**Table 1:** Estimation results of the degree of reporting delay

| | Estimated coefficient | $p$-value | 95%CI | |
| --- | --- | --- | --- | --- |
|  |  |  | Lower bound | Upper bound |
| 1/k | 17.28765 | .000 | 15.11999 | 19.4553 |
| 1/m | 1.009978 | .015 | 0.198866 | 1.821091 |
| 1/(km) | -18.19976 | .000 | -22.23761 | -14.16191 |
| constant | 0.0769113 | .745 | -0.3887593 | 0.542582 |

Note: The number of observations was 147. The coefficient of determination was 0.6603. $k$ denotes the number of days until the last reported day. $m$ denotes the difference between the publishing date in the past and the most recent publishing date. The dependent variable was the degree of reporting delay, which is defined as $X_{t-k-m}/t / X_{t-k-m}/t-m$ , where $X_{t-k}/t$ is the number of patients whose onset date was $t-k$ published on day $t$ . We used the number of patients by onset day published on 2, 5, 6, 10, 11 and 12 March 2020 by Ministry of Labour, Health and Welfare, Japan.

**Figure 2:**
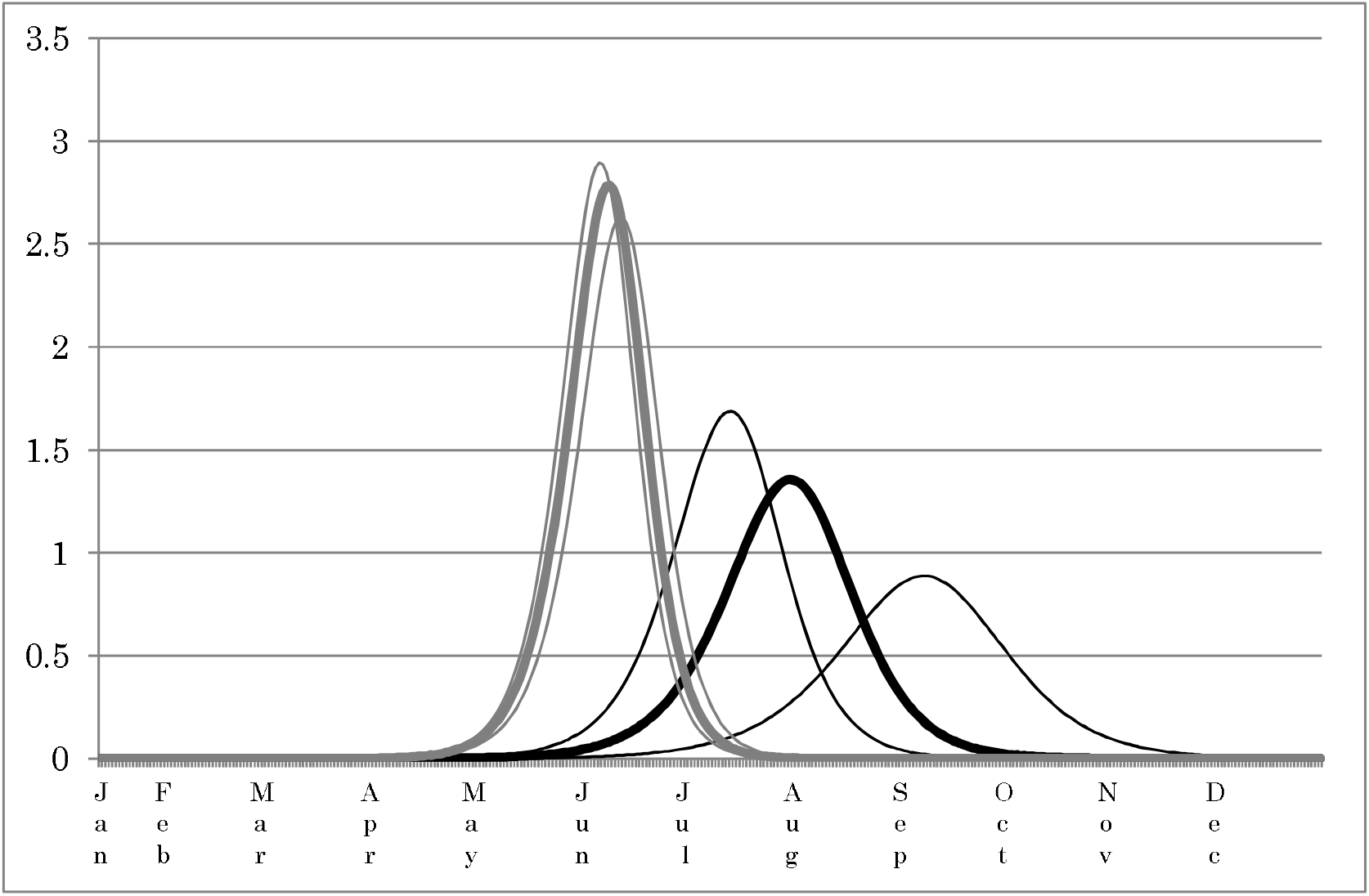
Predicted epidemic curve of COVID-19 patients with/without voluntary event cancellation in Japan and its 95% confidence interval. (million patients) Note: The black bold line represents the epidemic curve based on the estimated R_v_ with event cancellation countermeasures. Its 95% confidence interval (CI) is shown by the black thin lines. Gray lines represent the epidemic curves based on R_0_ without event cancellation countermeasures; its 95% CI is shown by gray thin lines.

The value of R_0_ before VEC introduced was estimated as 2.50. Its 95% CI was [2.43, 2.55]. However, R_v_ after VEC was introduced was estimated as 1.88; its 95% CI was [1.68,2.02].

Figure 3 depicts the entire predicted epidemic curve based on R_0_ and R_v_ and its 95% CI. The peak without VEC is estimated as being reached on 9 June with 2.78 million newly diagnosed patients with symptoms. The 95% CI peak dates are predicted as 7–12 June, with respective maximum numbers of patients with symptoms per day estimated respectively as [2.63, 2.89]. Under VEC, the peak occurred on 31 July [14 July, 9 September]. The maximum newly diagnosed patients with symptoms were 1.36[0.888, 1.69] million. In all, 81.5[80.3, 82.3] million patients showed symptoms under R_0_ and65.3[55.8, 70.3] million under R_v._ Particularly, the maximum number of inpatients was estimated as 3.63 [3.43, 3.76] million under R_0_ and 1.79 [1.18, 222] million under R_v._

## Discussion

We applied a simple SIR model including asymptomatic cases that had not been incorporated into the model to date. An earlier study [9] estimated R_0_ for COVID-19 as 2.24–3.58 in Wuhan. Our R_0_ obtained before VEC was similar.

However, R_v_ after VEC introduced was 35% reduction from R_0_. Actually, VEC can reduce the COVID-19 infectiousness by 35%. Even was after being reduced, it was higher than one. Therefore, VEC cannot contain the COVID-19 outbreak in Japan completely. The period of outbreak might be prolonged if VEC can be continued until the outbreak ceased. However, the maximum number of inpatients with VEC was lower than that without VEC by 70%. Therefore, we can expect mortality to be reduced because of the depletion of medical resources.

Nevertheless, the cost attributable to VEC must be considered. It is necessary to prove that the benefit attributable to reduction in patients and in mortality cases is greater than the cost of VEC. Such calculations remain as a subject for further research.

## Conclusion

Results demonstrated that VEC can reduce infectiousness of COVID-19 by 35%, although the R_0_ remains greater than one. The peak number of cases obtained in our simulation was decreased by about one third compared to the peak obtained without adoption of VEC. Moreover, the peak was delayed from occurring.

VEC had been extended until March 19 on March 10. We hope that the present study contribute government’s decision making for VEC. Results of the present study support the author opinions, but do not reflect any stance or policy of professionally affiliated bodies.

## Data Availability

All data are fully available without restriction.

https://www.mhlw.go.jp/stf/houdou/houdou_list_202001.html.

